# How robust are the results of one of the first positive trials exploring hydroxychloroquine for treatment of COVID-19?

**DOI:** 10.1101/2020.05.06.20093237

**Authors:** Ronald Chow, Sameer Elsayed, Michael Lock

**Author notes:** Correspondence: Ronald Chow, Schulich School of Medicine & Dentistry, University of Western Ontario, London, Ontario, Canada.

## Abstract

An outbreak of a novel human coronavirus infection emerged in Wuhan, China in December 2019. Two months later, the World Health Organization (WHO) announced SARS-CoV-2 as the name for the new virus and COVID-19 for the associated illness. On March 12, 2020, the WHO officially declared COVID-19 as a pandemic. The scientific community has raced to find effective therapeutic agents against the virus. Gautret *et al* 2020 is among one of the first purportedly positive trials of hydroxychloroquine for the treatment of COVID-19. However, it is imperative that a thorough analysis and understanding of trial data be undertaken prior to making claims about safety and efficacy. Our group assessed the statistical robustness of the trial using the Fragility Index (FI). The FI provides a numerical quantification of a clinical trial’s conclusions. The index is based on iterative statistical calculations to determine the minimum number of events within a trial that would theoretically need to change from positive to negative in order for the trial’s endpoint to convert from significant to non-significant; the higher the index, the more statistically robust the study results. For the Gautret *et al* trial, one endpoint had an FI of 1, two had indices of 2, and another had an index of 4. The primary endpoint of viral clearance on day 6 had an FI of 4. This indicates that if 4 events were to change from positive to negative, the conclusion of the trial would become mathematically non-significant. This index is comparable to many other published trials of established agents; the median FI across the reported literature appears to be 2. In conclusion, the trial results reported by Gautret *et al* are statistically robust, assuming that data quality is not compromised; however, the study was an open-label trial with non-homogenous groups, with analysis conducted per-protocol. Additionally, SARS-CoV-2 Reverse Transcriptase-PCR (RT-PCR) testing was not conducted in a systematic way amongst the two groups. Further analyses of this trial and future trials of antiviral agents with potential activity against SARS-CoV-2 should be performed with complementary epidemiologic and statistical techniques to determine whether the trial’s results are clinically important and/or should be explored in depth. Given the statistically robust results reported by Gautret *et* al, despite the study’s inherent methodological and analytical flaws, hydroxychloroquine should be studied as a potential agent against COVID-19 in larger clinical trials.

## Introduction

Coronaviruses primarily target the human respiratory system. Over the past two decades, these viruses were responsible for three epidemics of infection. Prior to 2019, the two coronavirus public health threats were caused by the Severe Acute Respiratory Syndrome (SARS-CoV) and Middle East Respiratory Syndrome (MERS-CoV) coronaviruses [1]. In December 2019, an outbreak of a novel coronavirus was reported in Wuhan, China [2] and subsequently rapidly spread around the world. By March 12, 2020, the World Health Organization (WHO) officially declared COVID-19 a pandemic. At the time of this writing in mid-April 2020, there are over 165,000 deaths worldwide [3]. Currently there is no effective treatment although the scientific community is racing to find possible active agents for COVID-19 [4]. One of the first purportedly positive trials was published by Gautret *et al* [5]. Earlier versions were released due the urgent need to provide data to health care providers as the virus was spreading rapidly across continents. This includes the publication in medRxiv [6] which had slightly different results denoted; as these were not peer reviewed, we only used data from the final peer reviewed publication in the International Journal of Antimicrobial Agents [5]. Gautret *et al* treated 20 cases with hydroxychloroquine in Marseille as the first large surge of cases reached the critical care departments of Europe; this study found that the drug was significantly associated with viral load reduction in COVID-19 patients [5]. After 6 days of treatment, 100%, 57.1% and 12.5% of the hydroxychloroquine plus azithromycin group, hydroxychloroquine alone group, and control group had negative viral RT-PCR tests, respectively. This observation was statistically significant at p<0.01.

Such findings are especially important during a public health crisis, where scientific discoveries need to be both rigorous and timely. Positive trial results could motivate the scientific community to use the agents in infected patients, or commit funding and clinical resources to explore this drug regimen further. However, it is important to pause for a moment to ensure that the purportedly positive trial results are indeed robust. Making inferences about this regimen without robust results may take away precious time and resources related to other promising therapies. Furthermore, the benefits of a new regimen need to be weighed against its potential harms; therefore, hydroxychloroquine use needs to be carefully weighed against potential harms such as prolonging of the QTc interval which can be life threatening.

There have been several critical appraisals of this trial, but appraisals are in many ways subjective and in this urgent and politicized subject, we sought to more quantitatively assess the value of this trial. To assess for robustness of a positive trial, a Fragility Index (FI) could be calculated. As described by Walsh *et al* [7], this index complements the p-value by informing the robustness of results. The index was developed by leaders in the field of evidence-based medicine, including Guyatt and Sackett, to provide information beyond the much criticized, but commonly reported p-value. The FI is most applicable and useful in trials with small numbers of events where the p-values can often erroneously suggest that a trial is positive; this may be the case for the small-sample trial published by Gautret *et al*. The index adds information beyond that provided by historically used measures such as *p*-value, number of events, confidence interval and sample size.

The aim of this paper is to determine the fragility of the trial by Gautret *et al*, one of the first purportedly positive trials exploring hydroxychloroquine for the treatment of COVID-19.

## Methods

### The Fragility Index

The FI is the minimum number of patients that needs to change from a negative to a positive event, counterfactually, in the non-favoured group for a study’s positive result to become negative/non-significant. The higher the FI, the more robust a positive trial’s results. The median FI of published positive trials is reported to be between 2 and 3 [8-11].

For example, in a trial where there are 10 patients in each the control and experimental arms and 5 patients with a positive event in the experimental arm, the results of 1 patient in the control arm needs to change from a negative to a positive event for a non-significant difference [Table 1]. As the number of events that needs to be changed is 1, the FI is therefore 1.

**Table 1.** Fragility Index Example

| Original Study |  |  |
| --- | --- | --- |
| $p = 0.033$ | Experimental Arm | Control Arm |
| Number of Events | 5 | 0 |
| Total Number of Patients | 10 | 10 |
| Fragile Study |  |  |
| $p = 0.141$ | Experimental Arm | Control Arm |
| Number of Events | 5 | 0 + 1 |
| Total Number of Patients | 10 | 10 |

### Statistical Methods

The number of patients with negative nasopharyngeal Reverse Transcriptase-PCR (RT-PCR) results among the number of patients per hydroxychloroquine and control arms, per day, were analyzed. To calculate the FI, the event number was incrementally increased by positive integer values until there was no statistical difference between the two trial arms. Fisher’s exact test was calculated for each counterfactual simulation, using Stata 15.

## Results

At day 3, 10 of 20 hydroxychloroquine patients had a virological cure, compared to 1 of 16 patients receiving placebo. A minimum of 2 additional control patients theoretically needed to have a virological cure in order to observe a nonsignificant difference. The FI for day 3 was 2 [Table 2]. The FI for virological cure by day 4 is 1, 2 for day 5, and 4 for day 6 [Tables 3-5].

**Table 2.** Fragility Index – Virological Cure (RT-PCR-negativity) by Day 3

| Original Study |  |  |
| --- | --- | --- |
| $p = 0.009$ | Hydroxychloroquine treated patients | Control patients |
| Number of Patients with Virological Cure | 10 | 1 |
| Total Number of Patients | 20 | 16 |
| Fragile Study |  |  |
| $p = 0.083$ | Hydroxychloroquine treated patients | Control patients |
| Number of Patients with Virological Cure | 10 | 1 + 2 |
| Total Number of Patients | 20 | 16 |

**Table 3.** Fragility Index – Virological Cure (RT-PCR negativity) by Day 4

| Original Study |  |  |
| --- | --- | --- |
| $p = 0.049$ | Hydroxychloroquine treated patients | Control patients |
| Number of Patients with Virological Cure | 12 | 4 |
| Total Number of Patients | 20 | 16 |
| Fragile Study |  |  |
| $p = 0.106$ | Hydroxychloroquine treated patients | Control patients |
| Number of Patients with Virological Cure | 12 | 4 + 1 |
| Total Number of Patients | 20 | 16 |

**Table 4.** Fragility Index – Virological Cure (RT-PCR negativity) by Day 5

| Original Study |  |  |
| --- | --- | --- |
| $p = 0.008$ | Hydroxychloroquine treated patients | Control patients |
| Number of Patients with Virological Cure | 13 | 3 |
| Total Number of Patients | 20 | 16 |
| Fragile Study |  |  |
| $p = 0.092$ | Hydroxychloroquine treated patients | Control patients |
| Number of Patients with Virological Cure | 13 | 3 + 2 |
| Total Number of Patients | 20 | 16 |

**Table 5.**
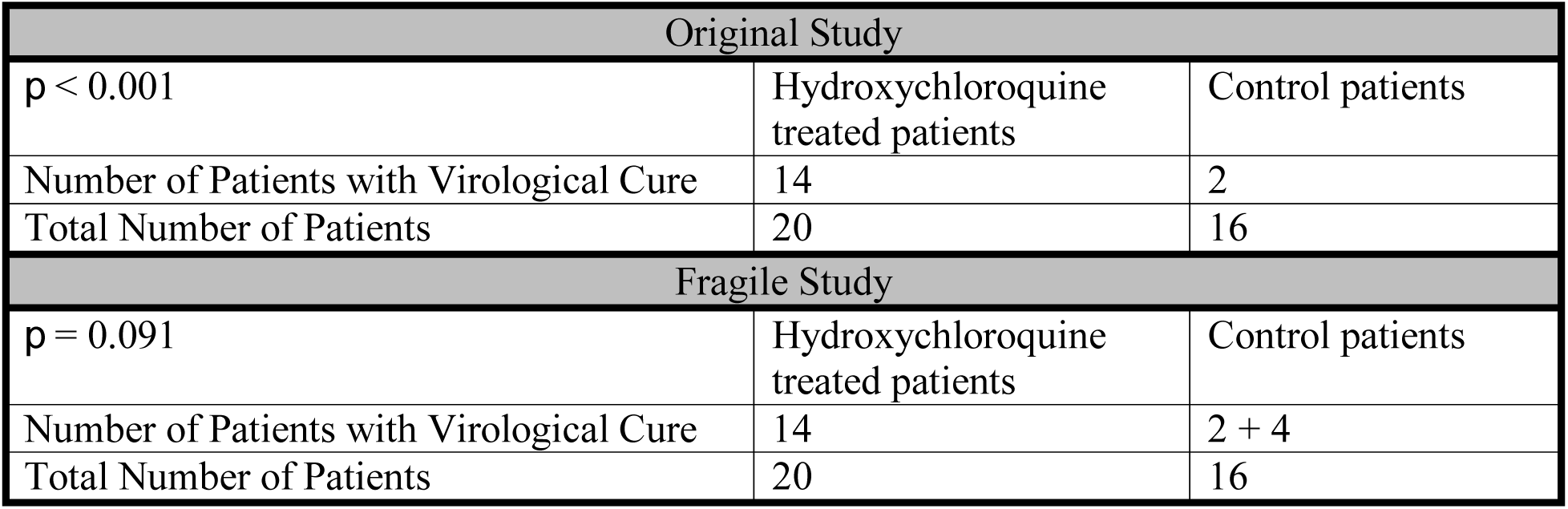
Fragility Index – Virological Cure (RT-PCR negativity) by Day 6

| Original Study |  |  |
| --- | --- | --- |
| $p < 0.001$ | Hydroxychloroquine treated patients | Control patients |
| Number of Patients with Virological Cure | 14 | 2 |
| Total Number of Patients | 20 | 16 |
| Fragile Study |  |  |
| $p = 0.091$ | Hydroxychloroquine treated patients | Control patients |
| Number of Patients with Virological Cure | 14 | 2 + 4 |
| Total Number of Patients | 20 | 16 |

## Discussion

This is the first study to quantitatively examine the results of Gautret *et al* from a statistical viewpoint. The FI ranged from 1 to 4. This would suggest that if as little as one event changed, the trial would have become statistically non-significant. It is unclear how this compares to other currently proposed agents against COVID-19 - two clinical trials of remdesivir in China were registered on ClinicalTrials.gov (NCT04252664 and NCT04257656) but suspended due to poor enrolment, with no results publicly available; other trials are still ongoing. This trial remains the only peer reviewed published COVID-19 drug treatment study.

Though the Gautret *et al* study has low indices, these indices are comparable to many other published trials. Evaniew *et al* reviewed 40 orthopedic spine surgery trials and reported a median FI of 2 [8]. Pediatric orthopedic trials have a median FI of 3, according to a review by Khormaee *et al* of 17 trials [11]. In the field of ophthalmology, the median index is 2 across 156 published trials [10]. Oncology trials are reported to have a median index of 2, according to a 36-trial review by Del Paggio and Tannock [12]. Perhaps the most pertinent assessment with respect to the COVID-19 trial by Gautret *et al* is a calculation of the FI in 57 critical care trials by Ridgeon *et al* that found a median FI of 2 [9]. The results of Gautret *et al* are as robust, statistically, as 50% of published trials [12].

Though reviewed in greater detail in other publications, a critical appraisal of the Gautret *et al* does highlight some aspects pertinent to the interpretation of the FI. Gautret *et al* have published essentially preliminary data from a practical and ongoing trial as they treated their first patients in Marseille, France. Thirty-six hospitalized patients with positive nasopharyngeal sample positive PCR results were included (20 treatment group and 16 untreated controls). Patients received hydroxychloroquine sulfate 200 mg three times per day for ten days, while six patients in the treatment group also received azithromycin. A comparison group was collected primarily in Marseille, Avignon, Briancon, and Nice. The primary endpoint was clearance of the virus on RT-PCR analysis of nasopharyngeal specimens on Day 6. Six patients were not included as they were lost to follow up. This is pertinent given that the FI indicates that the number of patients required to change the conclusion is 4. However, Walsh *et al* in their review of randomized trials published in high impact journals found that 52.9% of trials had higher loss to follow up levels than their Fragility Indices. The results of Gautret *et al* in this respect are similar to other published trials.

Also of note is the lack of systematic RT-PCR testing between the groups consequently time bias. The hydroxychloroquine group was, on average, more severely ill with COVID-19. Therefore, we would expect their RT-PCR tests to become negative quicker, even without treatment, due to a time bias. Furthermore, RT-PCR cannot even be used as a test of virological cure, as it measures viral RNA rather than the live virus. Given this methodological flaw, more trials are required to verify this purportedly positive trial.

The Fragility Index is only one method to provide insight into the value of a trial and has limitations. For example, the application to non-randomized trials is controversial as it lacks control of confounding variables. To further understand the results of Gautret *et al* and its applicability to future research and practice, focus should be spent on epidemiological aspects (i.e. trial design to control for confounder, inclusion/exclusion criteria, and choice of outcome), such as the analysis published by Kim *et al* [13].

In conclusion, the trial results reported by Gautret *et al* are statistically robust relative to other trials, but ideally need a higher Fragility Index and further investigation before the proposed regimen can be considered a viable treatment for COVID-19. Further analyses of the trial should be through the lens of epidemiology and clinical significance. Given the statistically robust results reported by Gautret *et* al, despite the study’s inherent methodological and analytical flaws, hydroxychloroquine should be studied as a potential agent against COVID-19 in larger clinical trials.

## Data Availability

N/A

## References

1. Rothan HA, Byrareddy SN. The epidemiology and pathogenesis of coronavirus disease (COVID-19) outbreak. Journal of Autoimmunity 2020; 109: 102433.

2. Lai CC, Shih TP, Ko WC, Tang HJ, Hsueh PR. Severe acute respiratory syndrome coronavirus 2 (SARS-CoV-2) and coronavirus disease-2019 (COVID-19): The epidemic and the challenges. International Journal of Antimicrobial Agents 2020; 55: 105924.

3. World Health Organization. Coronavirus disease 2019 (COVID-19). Situation Report – 89 [Internet]. Geneva: World Health Organization, 2020 Apr [cited 2020 Apr 18]. Available from: https://www.who.int/docs/default-source/coronaviruse/situation-reports/20200418-sitrep-89-covid-19.pdf?sfvrsn=3643dd382

4. Baden LR, Rubin EJ. Covid-19 - The Search for Effective Therapy. The New England Journal of Medicine 2020; doi: 10.1056/NEJMe2005477.

5. Gautret P, Lagier JC, Parola P et al. Hydroxychloroquine and azithromycin as a treatment of COVID-19: results of an open-label randomized clinical trial. International Journal of Antimicrobial Agents 2020; 105949.

6. Gautret P, Lagier JC, Parola P et al. Hydroxychloroquine and azithromycin as a treatment of COVID-19: results of an open-label non-randomized clinical trial. medRxiv 2020; doi: 10.1101/2020.03.16.20037135.

7. Walsh M, Srinathan SK, McAuley DF et al. The statistical significance of randomized controlled trial results is frequently fragile: a case for a Fragility Index. Journal of Clinical Epidemiology 2014; 67: 622–8.

8. Evaniew N, Files C, Smith C et al. The fragility of statistically significant findings from randomized trials in spine surgery: a systematic spine. Spine Journal 2015; 15: 2188–97.

9. Ridgeon EE, Young PJ, Bellomo R et al. The fragility index in multicenter randomized controlled critical care trials. Critical Care Medicine 2016; 44: 1278–84.

10. Shen C, Shamsudeen I, Farrokhyar F, Sabri K. Fragility of results in ophthalmology randomized controlled trials: a systematic review. Ophthalmology 2018; 125: 642–8.

11. Khormaee S, Choe J, Ruzbarsky J et al. The fragility of statistically significant results in pediatric orthopaedic randomized controlled trials as quantified by the fragility index: a systematic review. Journal of Pediatric Orthopaedics 2018; 38: e418–23.

12. Del Paggio JC, Tannock IF. The fragility of phase 3 trials supporting FDA-approved anticancer medicines: a retrospective analysis. Lancet Oncology 2019; 20: 1065–9.

13. Kim A, Sparks J, Liew J et al. A rush to judgement? Rapid reporting and dissemination of results and its consequences regarding the use of hydroxychloroquine for COVID-19. Annals of Internal Medicine 2020; doi: 10.7326/M20-1223.

